# Brief intensive social gaze training normalizes functional brain connectivity in boys with fragile X syndrome

**DOI:** 10.1101/2022.06.29.22277031

**Authors:** Manish Saggar, Jennifer L. Bruno, Scott S. Hall

## Abstract

Boys with fragile X syndrome (FXS), the leading known genetic cause of autism spectrum disorder (ASD), demonstrate significant impairments in social gaze and associated weaknesses in communication, social interaction, and other areas of adaptive functioning. Little is known, however, concerning the impact of behavioral treatments for these behaviors on functional brain connectivity in this population. As part of a larger study, boys with FXS (mean age 13.23 +/- 2.31 years) and comparison boys with ASD (mean age 12.15 +/- 2.76 years) received resting-state magnetic resonance imaging scans prior to and following social gaze training administered by a trained behavior therapist in our laboratory. Network-agnostic connectome-based predictive modeling (CPM) of pre-treatment RSFC data revealed a set of positive (FXS > ASD) and negative (FXS < ASD) edges that differentiated the groups significantly and consistently across all folds of cross-validation. Following administration of the brief training, the FXS and ASD groups demonstrated normalization of connectivity differences. The divergence in the spatial pattern of normalization response, based on functional connectivity differences pre-treatment, suggests a unique pattern of response to treatment in the FXS and ASD groups. These results support using connectome-based predictive modeling as an outcome measure in clinical trials.

## Introduction

Mutual eye-to-eye gaze (i.e., social gaze) is a key component of successful human communication, facilitating dyadic processes that form the basis of social interaction. Sensitivity to social gaze is present at birth (Farroni et al. 2002), and the development of social gaze over the first few years of life facilitates language learning (Çetinçelik et al. 2021). Impaired social gaze is a maladaptive feature associated with autism spectrum disorder (ASD) and is a particularly striking ‘hallmark’ feature of fragile X syndrome (FXS) (Hall et al. 2006). FXS is a rare inherited neurodevelopmental disorder that results from a mutation in the fragile X messenger ribonucleoprotein 1 (*FMR1)* gene (HGNC ID: 3775) and is the leading known genetic cause of ASD symptoms (Hagerman 2008). Given the known genetic cause, FXS can serve as a human model system for understanding the genetic and neurobiological underpinnings of autism symptoms, such as social gaze avoidance. This approach is supported by work from our lab and others demonstrating that FXS is associated with aberrant brain structural (Saggar et al. 2015; Bruno et al. 2016) and functional connectivity (Hall et al. 2013) as well as aberrant processing in response to social gaze (Bruno et al. 2014) and facial stimuli (Li et al. 2021). Further, research has shown that altered brain structure (Wolff et al. 2013) and brain function (Bruno et al. 2014) are related to ASD symptoms in individuals with FXS.

Treatments aimed at effectively normalizing aberrant social gaze behavior are of utmost importance given the social (Jones and Klin 2013; Cañigueral and Hamilton 2019) and educational (Çetinçelik et al. 2021) functions subserved by appropriate social gaze behavior. However, consideration of neural mechanisms underlying aberrant social gaze behavior is also critical. In FXS, aberrant social gaze is associated with physiological hyperarousal in terms of skin conductance (Williams et al. 2013), cortisol reactivity (Hessl et al. 2006), and pupillary reactivity (Farzin et al. 2009). Functional neuroimaging research using functional Magnetic Resonance Imaging (fMRI) (Bruno et al. 2014) and recently functional Near-Infrared Spectroscopy (fNIRS) (Li et al. 2021) has shown that neural systems underlying social gaze demonstrate plasticity, i.e., these systems change in response to repeated presentations over the course of several minutes. While previous studies showed sensitization to social gaze, likely due to enhanced social anxiety in children with FXS, plasticity indicates the capacity to change in response to gradual exposures in a controlled clinical setting. Consistent with this hypothesis, our previous proof of concept study demonstrated that a systematic behavioral skills training approach can be effective for teaching appropriate social gaze behavior in boys with FXS (Gannon et al. 2018). In particular, this approach utilized discrete trial instruction (DTI), a standardized teaching procedure that implements individualized instruction, well-defined steps, and a consistent rate of training trials to enhance learning (Smith 2001; Smith et al. 2009). Further, our broader intervention results utilizing DTI (Wilkinson, Britton & Hall, in press) indicated that brief behavioral skills training resulted in significant improvement in social gaze behavior for boys with FXS, demonstrated by decreased scores on an empirically validated parent-report questionnaire - the Eye Contact Avoidance Scale (ECAS) (Hall and Venema 2017). A symptom-matched comparison group of boys without FXS (who also had a diagnosis of ASD) did not show a change in ECAS scores following the same intervention. This result is potentially due to the unique characteristics of each disorder; individuals with FXS are sensitive to gaze initiation and may find eye contact aversive, whereas, in general, individuals with ASD may be insensitive to social gaze (Cohen et al. 1989).

In the present study, our goal was to examine changes in the functional organization of the brain associated with brief behavioral skills training in a subset of participants from the broader study (Wilkinson et al. in press). As reported previously, for adolescents/young adults with FXS, functional neuroimaging may represent a more sensitive outcome than a change in behavioral scores alone (Bruno et al. 2019). The higher sensitivity of neuroimaging markers (as opposed to behavioral markers) could be due to the fact that neural processes are intermediaries between *FMR1* gene mechanisms (i.e., reduced production of the fragile X messenger ribonucleoprotein (FMRP)) and social behavior. Furthermore, elucidation of brain mechanisms associated with treatment response is important for further refining behavioral interventions, including extending them to other populations.

Here we used resting-state functional connectivity (RSFC) to examine the functional reorganization of the brain following administration of a brief behavioral skills training package designed to promote appropriate social gaze. RSFC measures intrinsic, spontaneous co-fluctuations across brain regions not related to an explicit task (Kelly et al. 2012; Castellanos and Aoki 2016). RSFC is applicable across wide age ranges and levels of cognitive functioning (Castellanos and Aoki 2016), has good test/retest reliability (Thomason et al. 2011), and has been employed previously to examine differences in functional organization associated with FXS (Hall et al. 2013). To examine the changes in RSFC associated with the treatment, we first identified FXS-specific patterns of functional connectivity relative to an age and symptom-matched ASD comparison group of boys without FXS. We used a data-driven method initially developed to predict individual behavior from brain connectivity (connectome-based predictive modeling (CPM)) (Shen et al. 2017). An important advantage of CPM is that it allows one to define connectivity differences that are not limited to the classical functional brain network definitions. Finally, we examined intervention-related changes within the FXS-specific connectivity patterns.

## Methods

### Participants

Participants were recruited for the present study if they were male, aged 7 to 18 years, had a diagnosis of FXS or idiopathic ASD, and obtained a score of >/=30 points on the Eye Contact Avoidance Scale (ECAS), an empirically validated parent-report measure of social gaze avoidance (Hall and Venema 2017). Prior to visiting Stanford, participants were additionally screened with a questionnaire for the ability to comply with the image acquisition procedures. MRI preparation/desensitization involved having each participant review, at least twice, a 6-minute video via URL link showing a child having an MRI scan. The film includes footage of the MRI equipment and associated noises, descriptions of what the participant is required to do, and the images being acquired. Prior to their visit, participants were also required to listen to a 20-minute streaming audio via a URL link playing a collection of noises that occur during the course of a scan and that are associated with different pulse sequences the participant was likely to hear. The audio is narrated with information pertaining to the MRI experience and expectations for participant performance. Participants were also instructed to practice for the MRI at home by lying motionless on the floor under a chair with pillows placed firmly on either side of their head while listening to the audio track through headphones. Participants were invited to Stanford if parents reported that the participant could tolerate the MRI noises and could remain motionless for at least 10 minutes without moving their head.

Thirty-seven boys (16 FXS and 21 idiopathic ASD) met the inclusion criteria and completed the imaging procedures described below. FXS diagnosis was confirmed via genetic testing reports confirming aberrant methylation on the *FMR1* gene (> 200 CGG repeats). All individuals in the ASD comparison group obtained T-scores on the Social Responsiveness Scale, 2^nd^ Edition (SRS-2) (Constantino and Gruber 2012) above the cut-off of 60 for “autism risk,” with 80% of boys in this group also meeting the criteria for autism spectrum or autism on the Autism Diagnostic Observation Schedule, 2^nd^ Edition (ADOS- 2) (Gotham et al. 2009; Lord et al. 2012). The FXS and ASD groups were matched for sex, age, and baseline measures, including the Vineland Adaptive Behavior Scales, 2nd Edition (VABS-II) (Sparrow et al. 2005), the ADOS-2, SRS-2, and the ECAS (Hall and Venema 2017). Further inclusion/exclusion criteria and recruitment details are presented in Wilkinson, Britton, & Hall (in press), and participant flow is presented in Figure 1. Research was performed at the Stanford University School of Medicine and the Institutional Review Board approved all study procedures. Written, informed consent was obtained from a legal guardian for all participants.

**Figure 1.**
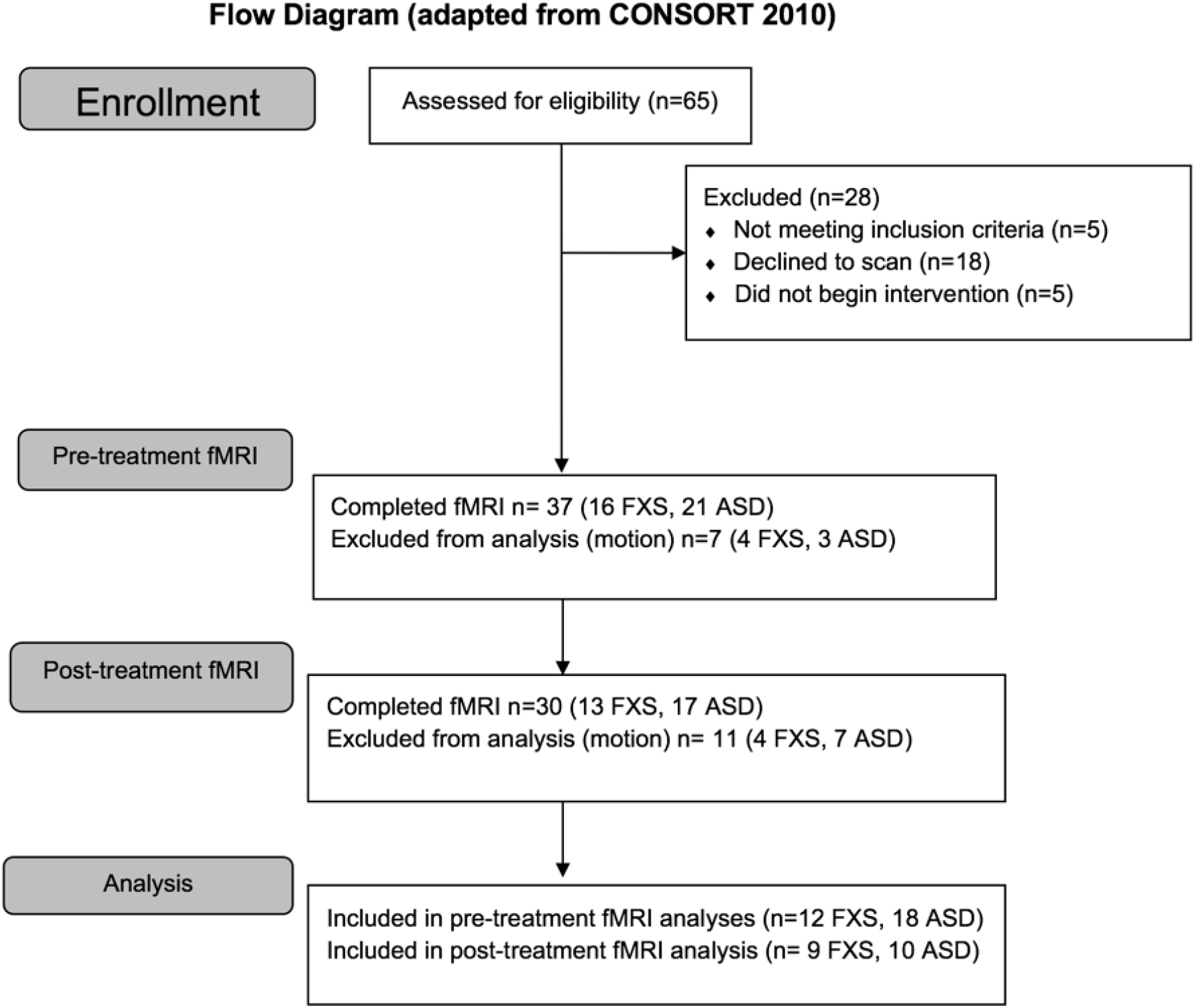
Study flow. ASD = autism spectrum disorder comparison group. fMRI = functional MRI

### Behavioral skills training

The intervention utilized a behavioral skills training approach to promote social gaze behavior (Gannon et al. 2018). Participants in each group received between 200 and 400 discrete training trials presented in four to eight 1-hour sessions (50 trials per session) over three days. During each treatment session, a trained behavior therapist reinforced longer durations of social gaze in discrete trials successively according to a percentile reinforcement schedule.

Sessions began by introducing a variety of deep breathing and progressive muscle relaxation exercises designed to decrease potential levels of physiological arousal (Gannon et al. 2018). Children were shown laminated cards containing icons for a *pufferfish, snowman, turtle, cat, batman*, and *lemon*, and were asked to choose three of the exercises and corresponding icons they would like to complete. The therapist then modeled the exercises, prompted the child to engage in the exercises, and provided verbal feedback when the child completed each exercise. These exercises generally took 5 minutes to complete.

Once the child had completed the relaxation exercises, the child sat in a chair directly facing the therapist. On *looking while listening* trials, the therapist stated, “I’m going to talk to you about something and I want you to look at my eyes while I talk to you”. Topics included the therapist telling a story (e.g., “Let me tell you about the time I went to…Let me tell you about the movie I saw this weekend…”). On *looking while speaking* trials, the therapist stated, “I want to learn more about you, I want you to tell me about the things you enjoy doing. Please look at my eyes while you talk to me.” On each trial, the therapist asked the participant questions about things they liked, activities they enjoyed doing, school (teacher, favorite subjects, friends, sports, etc.), their favorite foods, their family, where they live, etc. *Social gaze* was defined as the child orienting his head toward the therapist so that his eyes looked directly at the therapist’s face. If the duration of social gaze met the criterion for reinforcement according to the percentile schedule (see below), the therapist delivered verbal praise (e.g., “Good job!”), and awarded a token on a token board. If the participant did not engage in social gaze, or duration of social gaze did not meet criteria for reinforcement on a trial, increased verbal and gestural prompting was used to promote social gaze on the next trial. Reinforcement was delivered according to a percentile reinforcement schedule with the probability of reinforcing a criterion response (*w*), and the number of prior observations (*m*) to be included in the calculation set at .5 and 10 respectively (Galbicka 1994). Duration of social gaze therefore qualified for reinforcement on a particular trial if it exceeded 5 of the previous 10 response values (i.e., the median response value). This ensured that the rate of reinforcement was equivalent across participants. Once the participant had earned 10 tokens on the token board, the participant was allowed to play with a preferred item for 5 minutes. If the participant ended a session before earning all 10 tokens, the tokens carried over to the next session.

### Imaging procedures

All participants received MRI simulator training for 1 hour on Day 1 at Stanford (Barnea-Goraly et al. 2014). The MRI Simulator (Psychology Software Tools) includes a ∼60cm circular bore with cooling fans and lights, speakers for scanner noise production, and a movable table that can be operated from the control panel on the bore or by using the participant remote control. Accessories include a Mock head coil with rear-facing mirror, 30-inch flat panel LCD display, similar to the scanner display, and a Flock of Birds device for tracking head motion. The simulator protocol was designed to train participants to cooperate with the motion control requirements of MRI without the need for sedation. Behavioral techniques were used to counter anxiety experienced in association with the equipment and procedures. In brief, the process involved duplication and control of the salient stimuli in the imaging environment, gradual exposure to the equipment, personnel, and sensations involved in image acquisition, and reinforcement of the participant’s positive coping skills and efforts to inhibit body motion when instructed. These procedures were implemented with computer-assisted measurement and feedback for head motion over the 1-hour period.

After simulator training, participants completed imaging procedures on a GE 3.0 Tesla whole-body MR system (GE Medical Systems, Wilwaukee, WI) using a 32-channel head coil (Nova). Whole-brain functional images were collected during resting state using a T2-weighted multiband gradient echo pulse sequence and high-order shimming (echo time = 30 ms, repetition time=710 ms; acceleration factor = 6; flip angle=54°; field of view=22 cm x 22 cm; slice thickness = 2.4 mm, slice order = interleaved; approximate voxel size=2.4 mm^3^). Immediately after or before the resting-state scans, reference gradient-echo images were collected with opposing phase-encoding directions. Participants were instructed to relax and remain still in the scanner with their eyes closed during the 8 min resting-state scan. The first four frames were automatically discarded at the scanner, and the next ten frames were subsequently removed during preprocessing for scanner stabilization; thus, a total of 670 frames were available for subsequent data analysis. High-resolution T1-weighted (T1w) structural images were also collected during the same session to facilitate normalization to standard space (sagittal slices, repetition time 8.2 ms; echo time 3.2 ms; flip angle 12°; field of view 23 × 23 cm; matrix 256 × 256; 192 slices; voxel size = .9 mm^3^).

### Image preprocessing

Results included in this manuscript come from preprocessing performed using fMRIPrep 20.0.5 [(Esteban, Blair, et al. 2018; Esteban, Markiewicz, et al. 2018); RRID:SCR_016216], which is based on Nipype 1.4.2 [(Gorgolewski et al. 2011, 2018); RRID:SCR_002502]. The T1-weighted (T1w) image was corrected for intensity non-uniformity (INU) with N4BiasFieldCorrection (Tustison et al. 2010), distributed with ANTs 2.2.0 (Avants et al. 2008), and used as T1w-reference throughout the workflow. The T1w-reference was then skull-stripped with a Nipype implementation of the antsBrainExtraction.sh workflow (from ANTs), using OASIS30ANTs as the target template. Brain tissue segmentation of cerebrospinal fluid (CSF), white-matter (WM), and gray-matter (GM) was performed on the brain-extracted T1w using fast (FSL 5.0.9, RRID:SCR_002823, (Zhang et al. 2001)). Volume-based spatial normalization to two standard spaces (MNI152NLin2009cAsym, MNI152NLin6Asym) was performed through nonlinear registration with antsRegistration (ANTs 2.2.0), using brain-extracted versions of both T1w reference and the T1w template. The following templates were selected for spatial normalization: ICBM 152 Nonlinear Asymmetrical template version 2009c [(Fonov et al. 2009), RRID:SCR_008796; TemplateFlow ID: MNI152NLin2009cAsym], FSL’s MNI ICBM 152 non-linear 6th Generation Asymmetric Average Brain Stereotaxic Registration Model [(Evans et al. 2012), RRID:SCR_002823; TemplateFlow ID: MNI152NLin6Asym, (Zhang et al. 2001)].

The functional data were processed using the following steps. First, a reference volume and its skull-stripped version were generated using a custom methodology of fMRIPrep. A B0-nonuniformity map (or fieldmap) was estimated based on two (or more) echo-planar imaging (EPI) references with opposing phase-encoding directions with 3dQwarp (Cox and Hyde 1997) (AFNI 20160207). Based on the estimated susceptibility distortion, a corrected EPI (echo-planar imaging) reference was calculated for a more accurate co-registration with the anatomical reference. The BOLD reference was then co-registered to the T1w reference using flirt (FSL 5.0.9 (Jenkinson and Smith 2001)) with the boundary-based registration (Greve and Fischl 2009) cost-function. Co-registration was configured with nine degrees of freedom to account for distortions remaining in the BOLD reference. Head-motion parameters with respect to the BOLD reference (transformation matrices, and six corresponding rotation and translation parameters) are estimated before any spatiotemporal filtering using mcflirt (FSL 5.0.9, (Jenkinson et al. 2002)). The BOLD time-series (including slice-timing correction) were resampled onto their original, native space by applying a single, composite transform to correct for head motion and susceptibility distortions. The BOLD time series were resampled into standard space, generating a preprocessed BOLD run in MNI152NLin2009cAsym space. Several confounding time series were calculated based on the preprocessed BOLD: framewise displacement (FD), DVARS, and three region-wise global signals. FD and DVARS are calculated for each functional run, both using their implementations in Nipype (Power et al. 2014). The three global signals are extracted within the CSF, the WM, and the whole-brain masks (Jenkinson and Smith 2001; Greve and Fischl 2009, Jenkinson et al. 2002)

After fMRIPrep-based preprocessing, temporal masks were generated to flag motion-contaminated frames. Rigorous data preprocessing included flagging motion-contaminated frames in which framewise displacement (FD) was greater than 0.5 mm. For each such motion-contaminated frame, we also flagged one back and two forward frames as motion contaminated.

Following the construction of a temporal mask for censuring, the data were processed with the following steps: (i) demeaning and detrending, (ii), multiple regression, including time series from the: whole brain, CSF, and white matter signals, and motion regressors derived by Volterra expansion, where temporally masked data were ignored during beta estimation, (iii) interpolation across temporally masked frames using linear estimation of the values at censored frames so that continuous data can be passed through (iv) a second-order Butterworth band-pass filter (0.009 Hz < f < 0.08 Hz). The temporally masked (or censored) frames were then removed for further analysis. Lastly, we used group parcellation (Gordon et al. 2016) to create regional time series of preprocessed data into 333 cortical parcels. The Gordon parcellation is based on boundary maps defined using homogeneity of resting-state functional connectivity patterns. Participants were included in imaging analysis if at least 4 min of clean (or motion uncontaminated) data were available (Figure 1).

### Connectome-based predictive modeling

Connectome-based predictive modeling (CPM) was implemented using leave-one-out cross-validation (Shen et al. 2017). We first used the baseline (pre-treatment) data to find RSFC edges (between brain regions) that best differentiated the FXS and ASD groups while controlling for age and head motion (operationalized as the number of motion-contaminated frames). Non-parametric partial Spearman correlations of all RSFC edges were used to identify the most significant edges (p < 0.001) in the connectivity matrices that best differentiated the two groups. Specifically, for each iteration of the leave-one-out cross-validation, edges with a significant positive (FXS > ASD) and negative (ASD > FXS) correlation with p < 0.001 formed the positive and negative edge set, respectively, and were used as features to fit two linear models, i.e., polynomial regressions of degree 1. The resulting regression coefficients were then used to predict the group in the data of the remaining fold that was left out. The prediction based on the positive and negative edge sets was first evaluated within each fold by computing Spearman correlations between the predicted and true group membership. The positive and negative predictive cross-validated models were finally evaluated by correlating all predicted and true group memberships in the training set.

Next, we used the positive and negative edges that were consistently significant in each of the iterations of the leave-one-out cross-validation procedure applied to the training set to build a final linear regression model using the training data. The resulting consistent positive and negative edge sets were later used for the pre- vs post-treatment analysis, i.e., to examine intervention-related changes in RSFC in each group.

### Analysis of behavioral data

To examine potential changes in ECAS scores following treatment, we used modified Brinley plots (Blampied 2017). This graphical technique is useful for examining the effect of an intervention by depicting the data for each case as a coordinate pair across the two time points (e.g., baseline on the X-axis and post-treatment on the Y-axis) (McLay et al. 2021). Data points that lie below the 45° diagonal line (i.e., X = Y) are indicative of a therapeutic effect. Cohen’s *dav* Effect Size and Common Language Effect Size (CLES) were calculated (Lakens 2013). The CLES represents the probability (expressed as a percentage) that a score sampled at random from the post-treatment scores will be better than a score sampled at random from the pre-treatment score (McGraw and Wong 1992; Lakens 2013).

## Results

No significant differences were found between the groups on age, Vineland-II, ADOS-2, SRS-2 or ECAS scores (p’s > 0.05; Table 1), thus confirming group matching. Group matching was also confirmed for the subset of individuals who were included in pre-treatment MRI (12 FXS and 18 ASD, all p’s>0.10) and for the subset that were included in longitudinal (pre- and post-treatment) MRI (9 FXS and 10 ASD, all p’s>0.10).

**Table 1.**
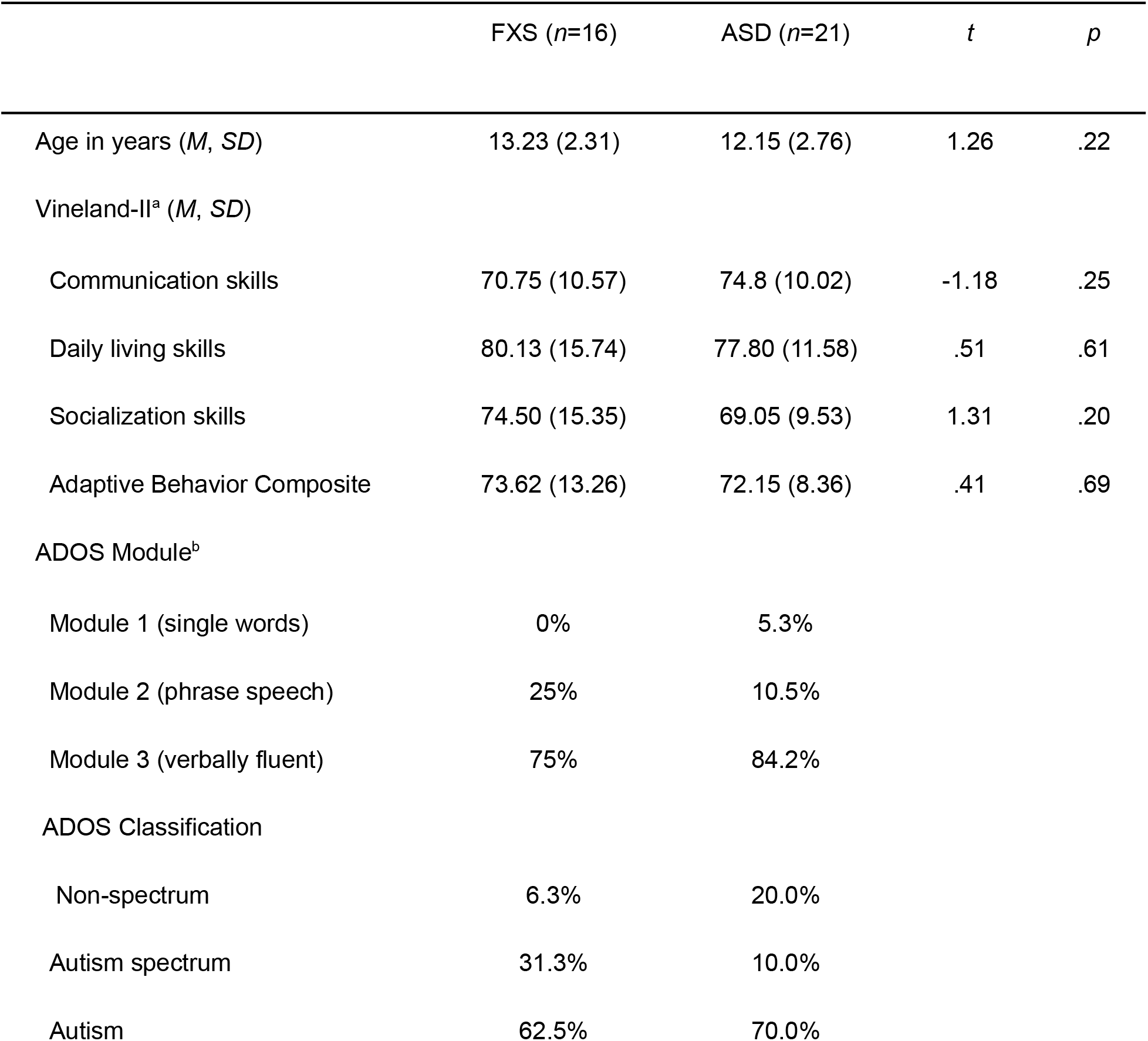
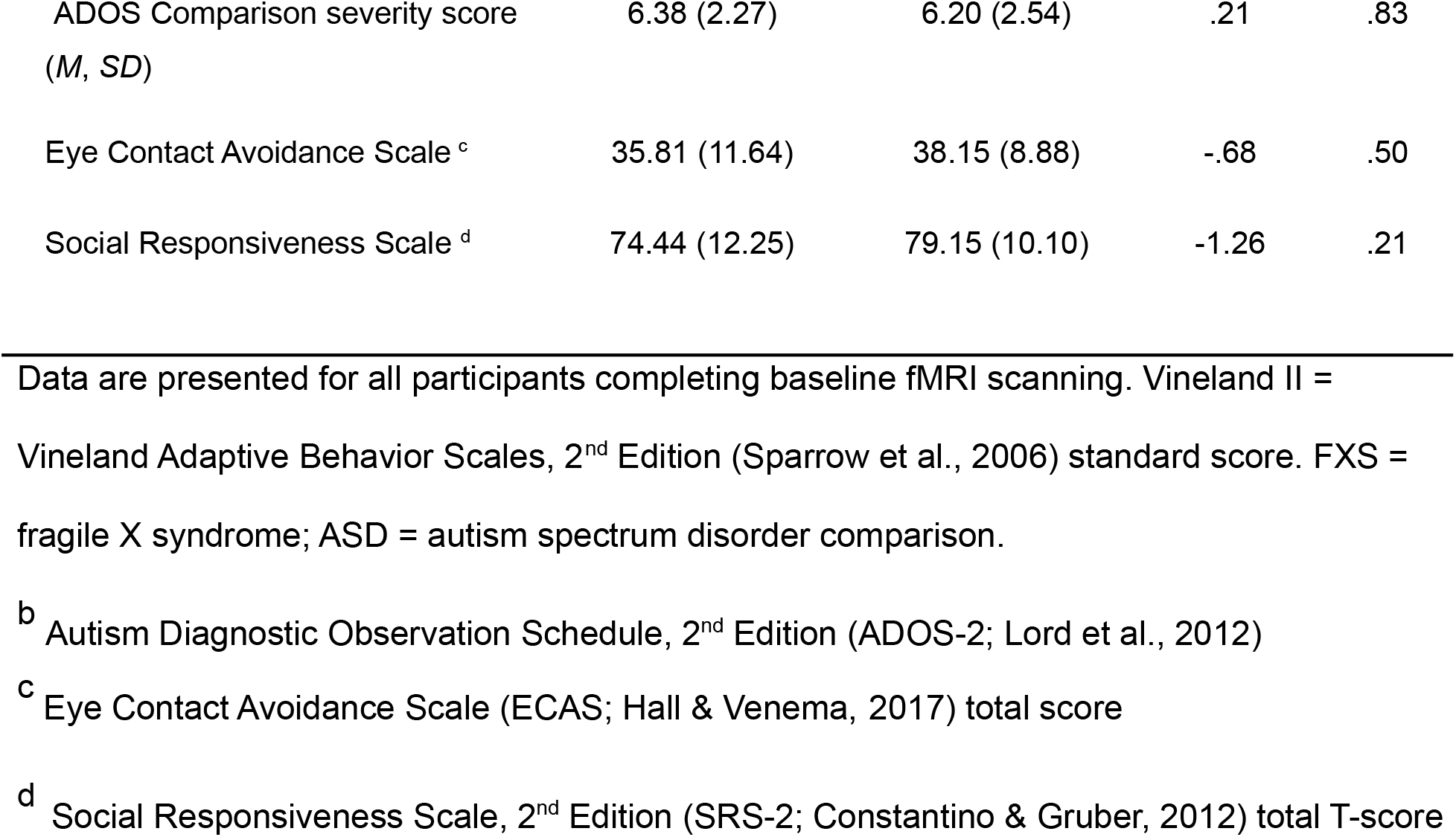
Participant characteristics at baseline.

### Group differences in RSFC at baseline (pre-treatment)

The CPM of pre-treatment RSFC data revealed a set of positive (FXS > ASD) and negative (FXS < ASD) edges that differentiated the groups significantly and consistently across all folds of cross-validation (Figure 2). Across all folds of cross-validation, the predicted group was significantly related to the actual group (while controlling for age and head motion) for both positive (Spearman ρ = 0.3695, p=0.0445) and negative edge sets (Spearman ρ = 0.4402, p=0.0149). Interestingly, our CPM-based approach revealed group differences in RSFC that were primarily *between* canonical resting-state networks, with a few exceptions of within-network differences (e.g., hyperconnectivity in FXS within the default mode network and hypoconnectivity in FXS within the fronto-parietal network). Primarily, FXS demonstrated higher between-network connectivity across the higher-order networks (e.g., between cingulo-opercular and fronto-parietal networks, and between fronto-parietal and dorsal attention networks). FXS also demonstrated lower (or hypo-) connectivity between default mode network and cingulo-opercular as well as fronto-parietal networks.

**Figure 2:**
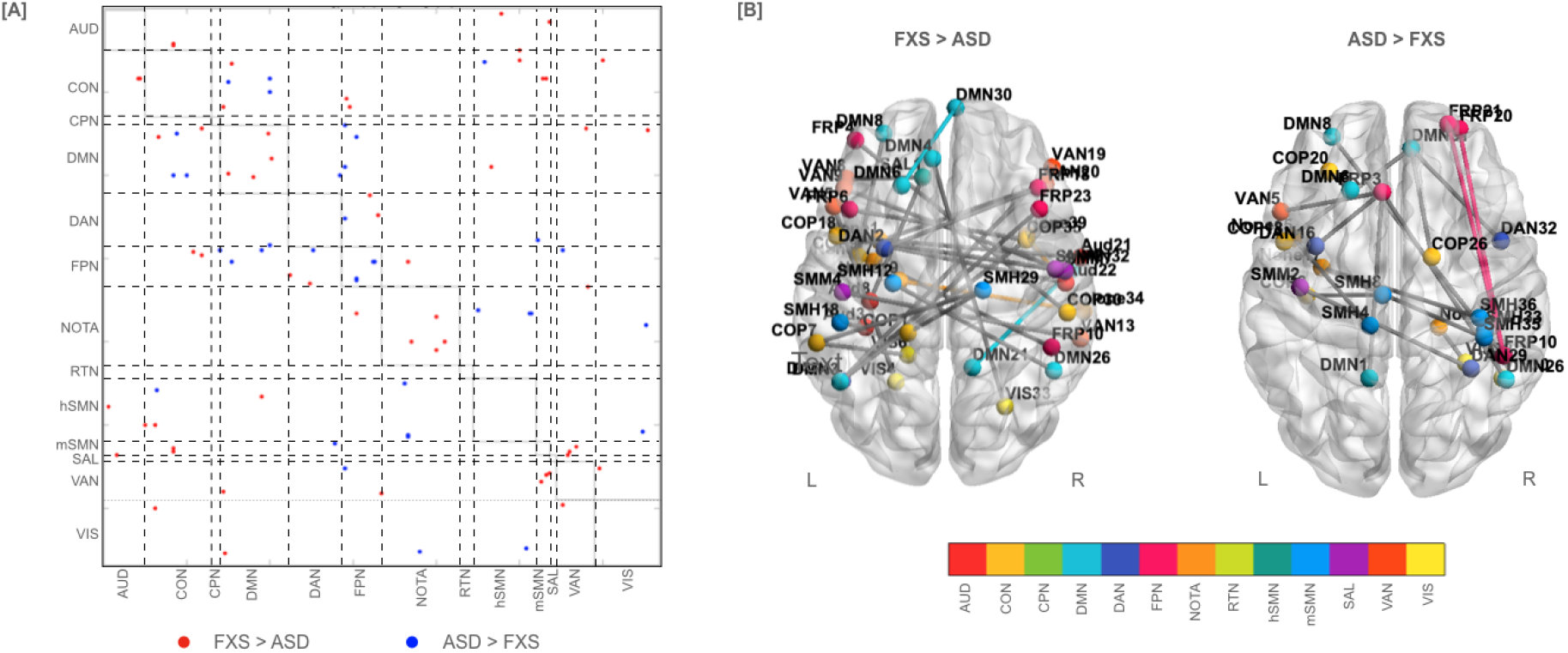
Group difference in RSFC at baseline (pre-treatment) as derived using the CPM approach. [A] Connectivity matrix (333 × 333 brain regions) showing consistent, across all folds of cross-validation, positive (FXS > ASD in red) and negative (ASD > FXS in blue) RSFC edges. [B] The consistent positive and negative RSFC edges are visualized on an average anatomical brain image to better depict edges that differentiate the two groups. Edges within a network are colored by the assigned network color. However, edges across (or between networks) are depicted using gray color. Interestingly, our CPM-based approach revealed group differences primarily in the between-network edges. Functional network abbreviations from the Gordon parcellation (Gordon et al. 2016): AUD: auditory; CON: cingulo-opercular; CPN: cingulo-parietal; DMN: default mode; DAN: dorsal attention; FPN: fronto-parietal; RTN: retrosplenial temporal; hSMN: hand somatomotor; mSMN: mouth somatomotor; SAL: salience; VAN: ventral attention; VIS: visual; NOTA: none of the above.

### Longitudinal changes in RSFC associated with behavioral skills training

Using repeated measures ANOVAs, while controlling for age and head movement, we assessed longitudinal changes in FXS-specific RSFC edge sets, as derived using the CPM-based approach at pre-treatment. Significant group x time effects were observed for both positive (FXS > ASD; F(1,15)=76.935, p<.001) and negative (ASD > FXS; F(1,15)=38.377, p<.001) RSFC edge sets. Post-hoc within-group pairwise t-tests (adjusted for multiple comparisons using Bonferroni correction) revealed significant changes in each group after treatment, such that hyperconnectivity within the FXS group was reduced, whereas the hypoconnectivity in the ASD group was also reduced post-treatment (Figure 3).

**Figure 3:**
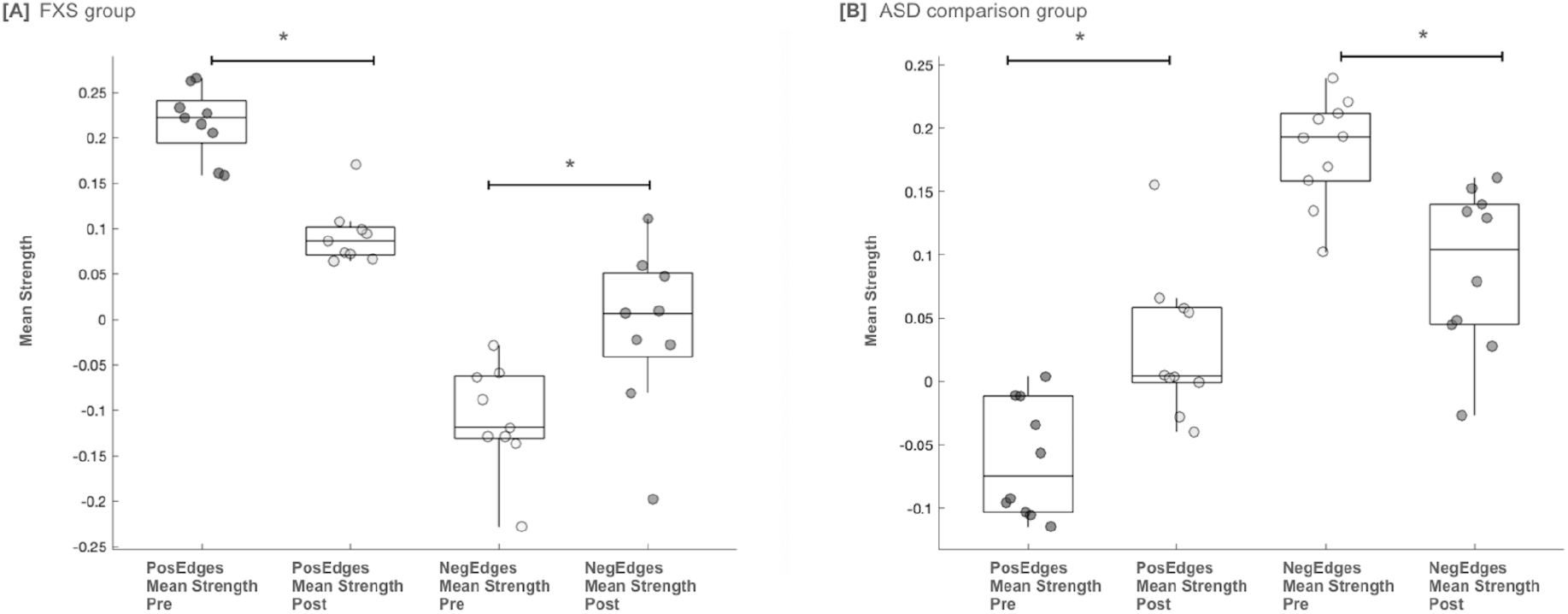
Longitudinal changes in positive (FXS > ASD) and negative (ASD > FXS) RSFC edge sets derived from pre-treatment data. Significant group x time interactions were observed for both positive and negative edge sets, such that a normalization of RSFC was observed post treatment for both groups. FXS = fragile X syndrome; ASD = autism spectrum disorder comparison group.; Pos = positive; Neg = negative

### Behavior changes associated with behavioral skills training

Figure 4 shows modified Brinley plots depicting total scores obtained on the ECAS at baseline and post-treatment for boys with FXS (left panel) and for boys with idiopathic ASD (right panel). The standardized mean difference effect size (*d*_*av*_) for ECAS scores from baseline to post-treatment was 1.03 for boys with FXS and .32 for boys with ASD (Figure 4). These data indicated that decreases in social gaze avoidance following the treatment probe were fairly large for boys with FXS and were more modest for boys with ASD. The Common Language Effect Size (CLES) was 86% for boys with FXS and 68% for boys with ASD.

**Figure 4:**
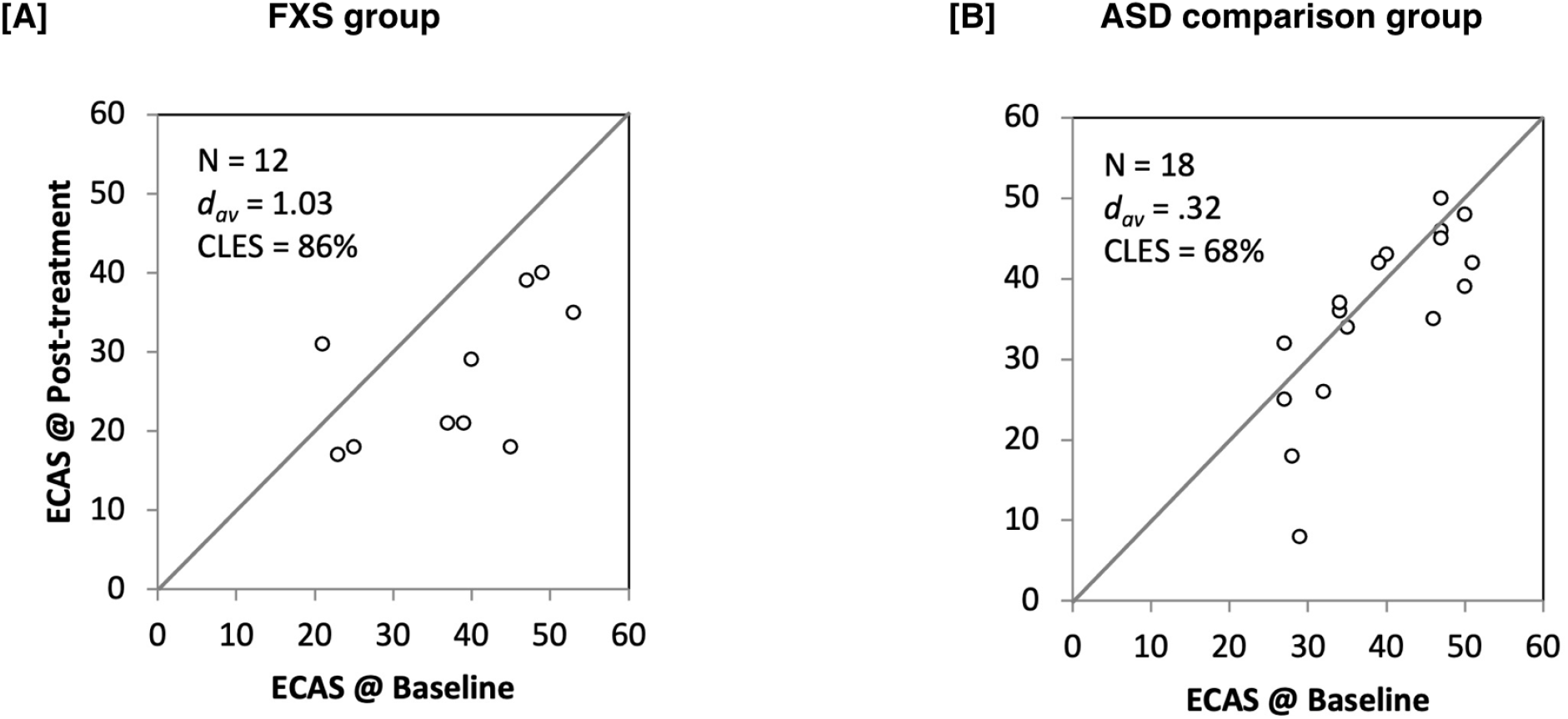
Visualization of change in social gaze behavior from baseline to post-intervention. Modified Brinley plots depict the effect of the intervention within each group. Data points that lie below the 45° diagonal line (i.e., X = Y) are indicative of a therapeutic effect. Effect sizes indicated that decreases in social gaze avoidance following the treatment probe were fairly large for boys with FXS and were more modest for boys with ASD. d_av_ **=** Cohen’s dav Effect Size; CLES = common language effect size; FXS = fragile X syndrome; ASD = autism spectrum disorder; ECAS = eye contact avoidance scale.

## Discussion

We provide the first evidence of changes in brain connectivity following administration of a brief behavioral skills training package targeted to promote social gaze behavior in boys with FXS and boys with ASD. We used resting-state functional MRI and connectome-based predictive modeling (CPM) to shed light on brain mechanisms associated with improvement in social gaze behavior. Within the FXS group, our results indicated widespread hyper- and hypo connectivity patterns at pre-treatment. Following treatment, we observed reorganization in functional connectivity patterns that indicated normalization of aberrant pretreatment connectivity differences in the FXS and ASD groups. This work specifies the neural mechanisms underlying social gaze training and has implications for future clinical trial research.

Pretreatment differences in brain connectivity revealed both positive edges (FXS>ASD) or hyperconnectivity and negative edges (FXS<ASD) or hypoconnectivity in boys with FXS for widespread regions of the brain. Only a handful of pre-treatment connectivity differences were localized within a canonical network, e.g., hyperconnectivity for FXS within default mode network and hypoconnectivity for FXS within the fronto-parietal network. In fact, most of the pretreatment connectivity differences involved connections between resting state networks, especially between the higher-order cognitive networks. Namely, the FXS group demonstrated hyperconnectivity between cingulo-opercular and fronto-parietal networks, and between dorsal attention and fronto-parietal networks. The FXS group also demonstrated hypoconnectivity between higher-order networks and the default mode network, namely between the frontoparietal network and the default mode network, and the cingulo opercular network and the default mode network. Hyperconnectivity may reflect a compensatory response to aberrant white matter development in association with the *FMR1* gene mutation, as evidenced by altered axon myelination in the mouse model (Pacey et al. 2013) and less efficient white matter connectivity in humans (Green et al. 2015). In contrast, hypoconnectivity may be related to aberrant gray and white matter structure found in individuals with FXS within areas of the fronto-parietal network (PFC, parietal regions) (Gothelf et al. 2008; Bray et al. 2011; Cohen et al. 2011). The combination of both hyper- and hypo-connectivity may be reflective of aberrant maturation of cortical networks as previous studies have indicated that within-network connectivity decreases while between-network connectivity strengthens in association with typical development (Rubinov and Sporns 2010; Lopez et al. 2020). Larger sample sizes and longitudinal work will be required to confirm these intriguing hypotheses. In summary, our pre-treatment results indicate a unique connectivity signature or “fingerprint” present in boys with FXS (Shen et al. 2017). Importantly, this “fingerprint” includes hypo- and hyperconnectivity within and between networks and extends previous work demonstrating only decreased, within network connectivity for a mixed group of male and female participants with FXS vs. a matched ASD comparison group (Hall et al. 2013).

Following behavioral skills training, the FXS group demonstrated a significant change in functional connectivity patterns within the set of edges that differentiated the FXS group at pre-treatment, i.e., the FXS-specific “fingerprint”. This result indicates a large-scale reorganization of functional connectivity patterns within and between canonical networks. Many of the edges showing change in parallel to improvement in social gaze involve at least one node linked to the social brain, including the ventral and medial prefrontal cortex, superior temporal gyrus, fusiform gyrus, cingulate gyrus, and amygdala (Adolphs 2009). Social gaze behavior is quite complex and involves many neural processes (Itier and Batty 2009; Senju and Johnson 2009; Carlin and Calder 2013). Thus, it is not surprising that the pattern of functional brain response to the treatment probe was equally complex. Furthermore, functional reorganization following the treatment probe indicated normalization of connectivity patterns for the FXS group. Specifically, the edges that demonstrated hyperconnectivity pre-treatment (positive edges) showed a decrease in connectivity strength post-treatment. Contrastingly, edges demonstrating hypoconnectivity pre-treatment (negative edges) showed an increase in connectivity strength post-treatment. Functional reorganization for the ASD group was in the same direction suggesting some shared mechanism in brain response following social skills training, namely normalization of functional connectivity differences pre-treatment. However, the divergence in the spatial pattern of normalization response, based on functional connectivity differences pre-treatment, suggests a unique pattern of response in the FXS and ASD groups. Divergence of spatial normalization patterns may be associated with group-specific motivation for gaze avoidance. Boys with FXS are more likely to be averse to social gaze due to social anxiety, whereas, in general, individuals with ASD demonstrate a lack of sensitivity to social gaze (Cohen et al. 1989).

Presently, the lack of a typically developing comparison group precludes us from understanding if the changes seen in either group are consistent with a trend toward typical functional brain connectivity patterns. Although we considered including a comparison group of typically developing individuals in our design, these individuals would not be matched on level of adaptive functioning, ASD symptomatology, or impairment in social gaze, thus any differences in RSFC would be confounded by those variables. We also note that only 37 of 60 (61.7%) participants who were eligible for scanning were able to tolerate and provide motion-free RSFC data at pre-treatment. The results may therefore not be representative of these clinical groups in general. Still, the normalization in functional connectivity response is an intriguing mechanism that warrants further study in FXS and in other clinical groups, in particular as a response to behavioral or other interventions.

Behavioral results indicated that decreases in social gaze avoidance, as quantified by the ECAS, following the treatment probe were fairly large for boys with FXS (*d*_*av*_ *=* 1.03) and were more modest for boys with ASD (*d*_*av*_ *=* .32). This is consistent with our larger intervention study, which demonstrated significant improvement in levels of social gaze for boys with FXS following behavioral skills training (Wilkinson, Britton, T & Hall, in press). Due to the limited sample size in the present fMRI study and the limited sensitivity of subjective behavioral outcomes in intervention research (Berry-Kravis et al. 2013; Bruno et al. 2019), we focused on effect sizes. We based this behavioral skills training on prior data showing that a carefully designed systematic behavioral skills training approach was effective for teaching appropriate social gaze behavior in boys with FXS (Gannon et al. 2018). Thus, it is not surprising that the treatment effect size on behavior was higher in boys with FXS in the present study. The significant change in brain functional connectivity we observed in the ASD group may indicate an initial response to the training that was not fully captured in the subjective ECAS outcome. Larger sample sizes, additional behavioral and neuroimaging outcome measures, and longer training durations will be important for confirming the findings of the present study and interpreting the utility of social gaze training for FXS in the longer term.

This is the first study to elucidate the brain mechanisms that underlie improvement in social gaze following behavioral skills training in FXS. We observed a reorganization of functional brain connectivity in parallel to improvement in social gaze, including reduced connectivity for edges that demonstrated pre-treatment hyperconnectivity and increased connectivity for edges that showed pre-treatment hypoconnectivity. Thus, the functional reorganization associated with improvement in social gaze can be viewed as normalization of pre-treatment FXS-specific brain connectivity differences. These results support using functional neuroimaging and connectome-based predictive modeling as outcome measures in future clinical trials.

## Data Availability

The final dataset will be stripped of all identifiers and made available to qualified investigators upon request.

## Acknowledgments

We thank the families who volunteered to participate in this study. We also wish to acknowledge Tobias Britton and Ellen Wilkinson for their help with recruitment and data collection.

